# Reducing the carbon footprint of inhaler prescriptions at an Australian tertiary health service: a modelling exercise

**DOI:** 10.1101/2025.07.12.25331451

**Authors:** Joshua VB Taverner, Lata Jayaram

**Author notes:** **Corresponding Author** Dr. Joshua Taverner Western Health, 176 Furlong Road, St Albans VIC 3021, Australia.

## Abstract

**Background:** Climate change poses significant health risks, particularly for respiratory patients. Healthcare systems contribute substantially to greenhouse gas emissions, with inhalers representing a notable source. Metered dose inhalers contain hydrofluorocarbons with global warming potentials 1,300-3,350 times that of carbon dioxide, while dry powder inhalers and soft mist inhalers have negligible emissions. Despite guidelines recommending environmentally friendly alternatives, limited data exists on Australian hospital discharge prescribing patterns and their environmental impact.

**Methods:** We conducted a retrospective descriptive study of all inhaler prescriptions at discharge from Western Health, Victoria, from January 1 to December 31, 2023. De-identified prescription data were extracted from electronic medical records without exclusions. We calculated total prescriptions by device type and medication class, estimated carbon emissions using published propellant data, and modelled potential emission reductions from switching metered dose inhalers to dry powder inhalers where clinically appropriate. Scenarios included 10%, 50%, 85%, and 100% conversion rates.

**Results:** Of 16,769 inhaler prescriptions, 71.87% were metered dose inhalers, 22.11% dry powder inhalers, and 6.02% soft mist inhalers. Short-acting medications comprised 52.7% of prescriptions, with 99.0% of short-acting beta-agonists prescribed as metered-dose inhalers. Total estimated carbon emissions were 296.35 tonnes carbon dioxide equivalent, with metered-dose inhalers contributing 98.79% (292.8 tonnes). Short-acting beta-agonists alone accounted for 64.22% of total emissions. If all clinically appropriate metered-dose inhalers were switched to dry powder inhalers, emissions would decrease by 267.79 tonnes (90.36% reduction). More modest changes would yield substantial benefits: 85% conversion (227.62 tonnes reduction), 50% conversion (133.89 tonnes), and 10% conversion (26.78 tonnes).

**Conclusions:** This study addresses a critical knowledge gap by providing the first comprehensive analysis of hospital discharge inhaler prescriptions and their environmental impact in Australia. Metered-dose inhaler prescriptions at hospital discharge contribute disproportionately to healthcare carbon emissions, with the majority coming from short-acting medications. Substantial emission reductions are achievable through the preferential prescribing of dry powder inhalers where clinically appropriate. Hospital discharge represents a critical intervention point, as these prescriptions often template ongoing community prescribing. These findings provide essential baseline data for implementing Australia’s National Health and Climate Strategy and achieving significant healthcare emission reductions while maintaining quality care.

## Article

### Background

Climate change poses unprecedented threats to global health, with respiratory patients particularly vulnerable to increased pollen loads, thunderstorm asthma events, and worsening air quality (1)(2). Greenhouse gas emissions trap heat in the Earth’s atmosphere, leading to global warming and climate change, which in turn creates these health risks. Healthcare systems themselves contribute considerably to this problem through their greenhouse gas emissions. The Australian healthcare system contributes as much as 7% of Australia’s greenhouse gas emissions (2), significantly higher than the 4.4% global average (3). Inhalers used to treat respiratory diseases such as asthma or chronic obstructive pulmonary disease (COPD) have been shown to be responsible for 3% of healthcare sector emissions in the comparable United Kingdom (UK) National Health Service (NHS)(4).

Metered dose inhalers (MDIs) are the most prescribed device type and contain hydrofluorocarbons (HFCs). Hydrofluorocarbons have a considerable global warming potential (GWP), which measures how much heat a gas traps compared to carbon dioxide (CO^2^) over a 100-year period, thereby contributing to climate change. The HFCs commonly used in MDIs include HFC-134a and HFC-227ea, which have GWP values of 1,300 and 3,350 times that of carbon dioxide, respectively. By contrast, Dry Powder Inhalers (DPIs) and Soft Mist Inhalers (SMIs) do not contain hydrofluorocarbon propellants and contribute negligible greenhouse gas emissions (4), hereafter referred to as ‘carbon emissions’. Most medications administered via MDI are also available via DPI in Australia, or there are medicines of the same class and efficacy available via DPI. With some exceptions, almost all patients can be safely prescribed an environmentally friendly inhaler with equal or superior outcomes than would be achieved via MDI (5).

Changing inhaler prescriptions to low-carbon alternatives aligns with local guidelines from the Thoracic Society of Australia and New Zealand (TSANZ) (6) and international guidelines such as those by the British Thoracic Society (1,2). In Australia, the National Health and Climate Strategy, released in December 2023, aims to reduce the environmental impact of healthcare. The reduction in carbon emissions from inhaler use has been identified as an area for improvement (7). Many patients are motivated to consider environmental impact when choosing their inhaler, and a change in inhalers can reduce carbon emissions comparable to switching from a gasoline to a hybrid car or adopting a plant-based diet(6)(8).

Despite this, there is limited published data on inhaler device prescriptions in Australia, especially upon hospital discharge. To address this knowledge gap, this study aims to quantify the proportion of each inhaler device prescribed on discharge from a large metropolitan hospital in Melbourne and to estimate the carbon impact from the prescribed inhalers using previously published data. By calculating potential carbon savings from reducing MDI prescriptions, this study will estimate the environmental benefits of adopting more low-carbon inhaler prescribing practices at discharge.

### Methods

We performed a retrospective descriptive study of inhaler prescriptions on discharge from Western Health, Victoria, from January 1 to December 31, 2023. Western Health is a large metropolitan health service comprising two major hospitals and several smaller facilities in western Melbourne, serving a diverse population with three emergency departments and specialised units, including respiratory, general medicine, and geriatrics. After obtaining ethics approval from the Western Health ethics committee (Ethics number 7326409), we obtained de-identified inhaler prescriptions on discharge for every patient with an inpatient stay at Western Health in the year 2023 via data collection from the electronic medical record. We did not exclude any patients or prescriptions from this study. Clinical details were unavailable as patient records were not accessed. The data was stored in a password-protected Excel file on the hospital server, and calculations and analysis were performed using Microsoft Excel and figures created with HTML.

We calculated the total prescription of each inhaled medication by device type, assuming each prescription was filled once following hospital discharge, without assuming repeat prescriptions or continued prescriptions within the community, as we could not utilise community pharmacy dispensing data. We calculated the total carbon emissions for each inhaler using carbon footprint estimates by Tirumalasetty et al., which were derived from propellant weight, global warming potential values, and lifecycle analysis data. However, these estimates carry uncertainty due to reliance on material safety data sheets rather than direct measurement and extrapolation of manufacturing emissions (9). We then calculated the total carbon emission for each class of medication and each combination of classes. Preventer inhalers typically contain a one-month supply of medication, and Australian hospitals are responsible for prescribing medications to cover the month following discharge. Therefore, our analysis represents carbon emissions from inhalers for the first month post-discharge. This descriptive study did not require statistical testing as we analysed all discharge prescriptions rather than a sample.

For each MDI prescribed, we identified environmentally friendly DPI alternatives with substantially lower carbon emissions, if possible. Where available, we identified a DPI with the same medication(s) or, if not possible, a DPI with medication(s) from the same class and of equivalent efficacy. We then calculated the estimated carbon emissions if 10%, 50%, or 85% or all MDIs were changed to DPIs. We selected these estimates to represent a modest percentage change (10%), a substantial change (50%), or a change that would align Australian prescriptions with those in Sweden (85%). The potential carbon savings were then calculated to provide an estimate for how changing hospital prescriptions would directly reduce the environmental impact of inhalers.

### Results

A total of 16,769 inhalers were prescribed at Western Health in 2023 (Table 1), with short-acting agents comprising over half (52.7%) of all prescriptions. Table 1 provides a detailed breakdown of prescriptions by medication class and device type. Short-acting medications dominated prescribing patterns, with short-acting beta agonists (SABAs) representing the largest single category at 7,610 prescriptions (45.38%) and short-acting muscarinic antagonists (SAMAs) adding another 1,235 prescriptions (7.36%). Single-agent preventers made up a small proportion of prescriptions, with 26 (0.16%) long-acting beta-agonists (LABAs), 1,491(8.89%) long-acting muscarinic antagonists (LAMAs), and 756 (4.51%) inhaled corticosteroids (ICS). Combination preventers made up the remaining inhaler prescriptions with 414 (2.47%) long-acting beta-agonist and long-acting muscarinic antagonist (LABA/LAMA) prescriptions, 3,783 (22.56%) inhaled corticosteroid and long-acting beta agonist (ICS/ LABA) prescriptions and 1454 (8.67%) triple therapy prescriptions containing inhaled corticosteroids, long-acting beta-agonist and long-acting muscarinic antagonist (ICS/ LABA/ LAMA). Overall, short-acting medications accounted for most of the total prescription volume, while combination ICS/LABA medications represented the largest single category of preventer prescriptions.

**Table 1:** Distribution of Inhaler Prescriptions by Medication Class and Device Type.

| Medication Class | MDI | DPI | SMI | Total | Percentage |
| --- | --- | --- | --- | --- | --- |
| Short-Acting Beta-Agonist (SABA) | 7,534 | 76 | 0 | 7,610 | 45.38% |
| Short-Acting Muscarinic Antagonist (SAMA) | 1,235 | 0 | 0 | 1,235 | 7.36% |
| Long-Acting Beta-Agonist (LABA) | 0 | 26 | 0 | 26 | 0.16% |
| Long-Acting Muscarinic Antagonist (LAMA) | 0 | 717 | 774 | 1,491 | 8.89% |
| LABA/LAMA Combination | 0 | 179 | 235 | 414 | 2.47% |
| Inhaled Corticosteroid (ICS) | 484 | 272 | 0 | 756 | 4.51% |
| ICS/LABA Combination | 2,319 | 1,464 | 0 | 3,783 | 22.56% |
| ICS/LABA/LAMA Triple Therapy | 480 | 974 | 0 | 1,454 | 8.67% |
| **TOTAL** | **12,052** | **3,708** | **1,009** | **16,769** | **100.00%** |
| **Percentage by Device** | **71.87%** | **22.11%** | **6.02%** | **100.00%** | **—** |

**Legend:** SABA = Short-Acting Beta-Agonist; SAMA = Short-Acting Muscarinic Antagonist; LABA = Long-Acting Beta-Agonist; LAMA = Long-Acting Muscarinic Antagonist; ICS = Inhaled Corticosteroid; MDI = Metered Dose Inhaler; DPI = Dry Powder Inhaler; SMI = Soft Mist Inhaler. Data represents inhaler prescriptions dispensed at a tertiary hospital over 12-month study period.

In terms of device types, most prescriptions were MDIs, with 12,052 (71.87%) MDI prescriptions, 3,708 (22.11%) DPI prescriptions, and 1,009 (6.02%) SMI prescriptions. These findings are consistent with a recent study from the nearby Royal Melbourne Hospital by Cushnahan et al, which reported that 79% of inpatient inhaler prescriptions were MDIs over a 12-month period, closely matching our 71.87% proportion (8). As shown in Figure 1, the distribution of device types varied considerably across medication classes. Table 1 demonstrates that short-acting medicines were almost entirely prescribed as MDIs, with 99.0% of SABAs and 100% of SAMAs (which are only available as MDIs) dispensed in this form. Single-agent preventers were considerably more likely to be prescribed via an environmentally friendly device. This variation in device selection across medication classes is clearly illustrated in Figure 1. Single-agent LABA or LAMAs are only available via environmentally friendly options and so were all prescribed via non-MDI form, with 26 (100%) LABAs prescribed as DPIs, 717 of 1,491 (48.09%) LAMA prescriptions prescribed as DPIs, and 774 (51.91%) as SMIs. Inhaled corticosteroid prescriptions were mixed, with 484 (64.02%) being prescribed as MDIs and 272 (35.98%) being prescribed as DPIs. Combination LAMA/ LABA medications are only available in environmentally friendly devices with 179 (43.24%) prescribed as DPIs and 235 (56.76%) prescribed as SMIs. Combination medications that contain a corticosteroid are available in either MDI or DPI form. ICS/ LABA medications were prescribed primarily in MDI form, with 2,319 (61.30%) MDI prescriptions compared to 1,464 (38.70%) DPI prescriptions. Contrastingly, triple therapy prescriptions were primarily prescribed in DPI form, with 974 (66.99%) DPI prescriptions compared to 480 (33.01%) MDIs. Device selection varied markedly by medication class, with short-acting medications almost exclusively prescribed as MDIs, single-agent preventers predominantly prescribed as DPIs or SMIs, and combination preventers showing significant variation in the proportion prescribed as MDIs versus other devices.

**Figure 1.**
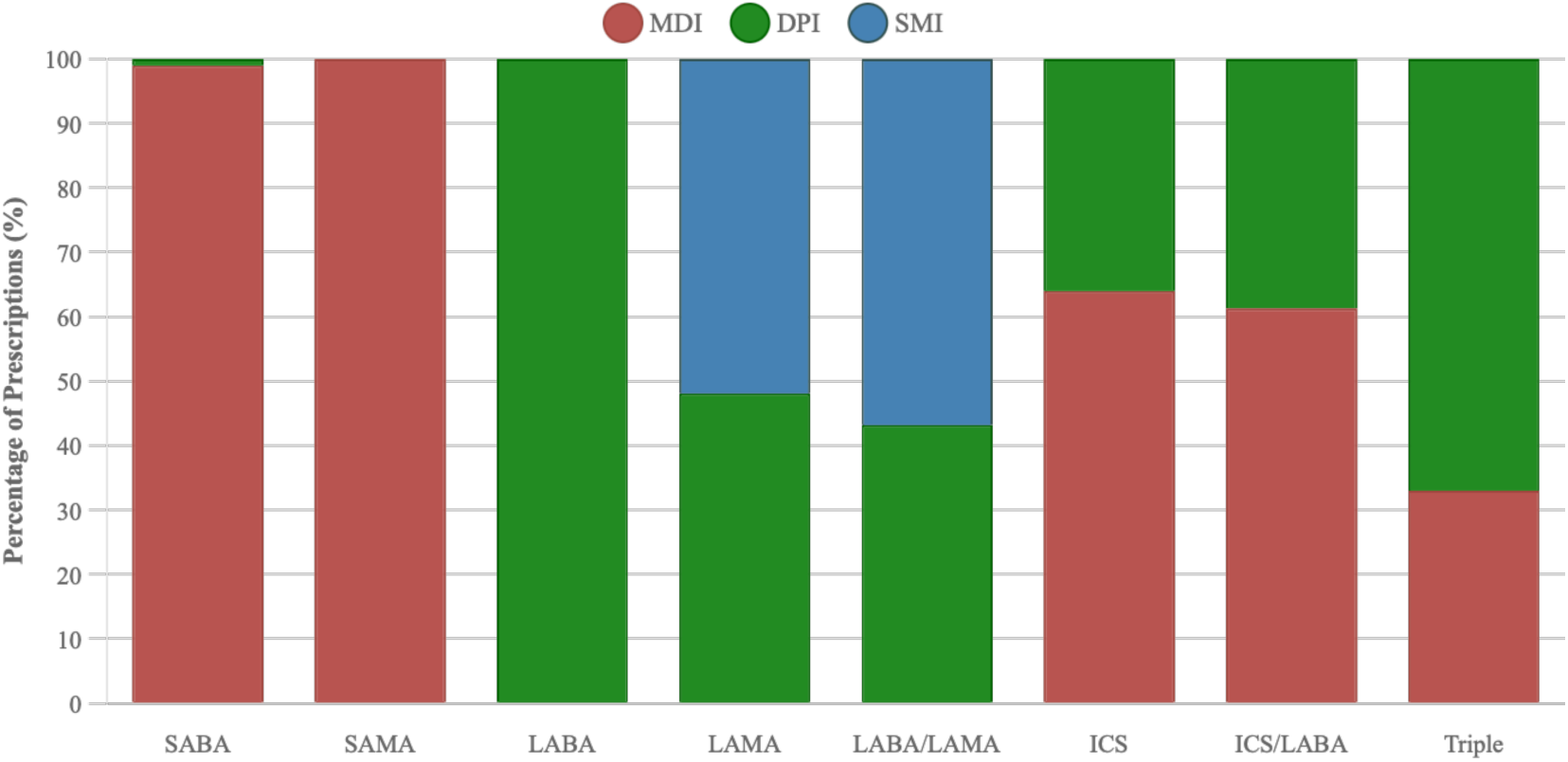
Medication class prescriptions by device type. Proportion of device types prescribed within each medication class at Western Health in 2023. Short-acting medications were almost exclusively prescribed as MDIs (99.0% of SABAs, 100% of SAMAs). Preventive medications showed greater variation, with ICS/LABA combinations predominantly prescribed as MDIs (61.3% vs 38.7% DPIs), while triple therapy inhalers were more commonly prescribed as DPIs (67.0% vs 33.0% MDIs). Single-agent LABAs and LAMAs are only available as DPIs/SMIs. MDI = metered-dose inhaler; DPI = dry powder inhaler; SMI = soft mist inhaler.

**Figure 2.**
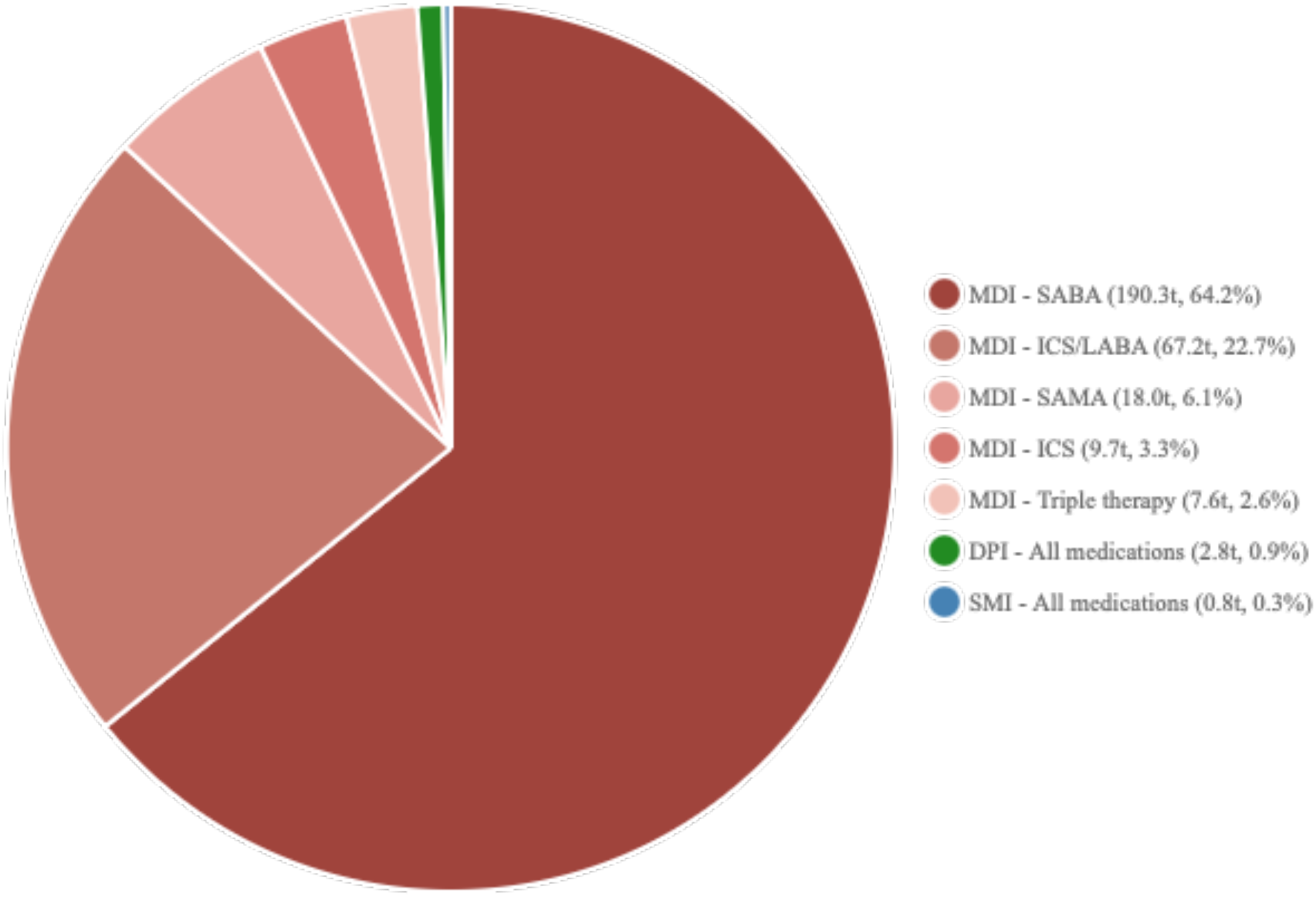
Carbon Emissions by inhaler (Tonnes) Total carbon dioxide equivalent emissions (296.4 tonnes) by device type and medication class. MDI-SABA contributed 64.2% of emissions (190.3 tonnes) despite representing 45.4% of prescriptions. MDI-ICS/LABA inhalers contributed 22.7% of emissions (67.2 tonnes) from only 13.8% of prescriptions. All DPI and SMI devices combined contributed 1.2% of total emissions (3.6 tonnes). SAMA = short-acting muscarinic antagonist; ICS = inhaled corticosteroid; LABA = long-acting beta-agonist.

Using carbon emission estimates by Tirumalasetty et al (9), the environmental impact varied dramatically by device type, ranging from 588.5g per device for the lowest-impact option (Symbicort Turbuhaler) to 36,500g per device for the highest-impact MDIs (Symbicort Rapihaler and Flutiform) due to the potent HFA 227ea propellant. An estimated 296.35 tonnes of carbon emissions were released due to inhaler prescriptions as detailed in Table 2. The emissions were primarily due to MDI prescriptions (Table 3), with 292.8 tonnes (98.79%) equivalent of CO^2^ from MDIs compared to 2.8 tonnes (0.94%) from DPIs and 0.78 tonnes (0.26%) from soft mist inhalers. Most of the 292.8 tonnes of carbon emissions attributed to MDIs were from SABAs and SAMA prescriptions at 190.3 tonnes (65.00%) and 18.0 tonnes (6.15%), respectively. The Symbicort Rapihaler and Flutiform are both ICS/ LABA devices that utilise HFC 227ea and are consequently responsible for a disproportionate amount of MDI carbon emissions at 67.2 tonnes (22.96%) despite making up only 2319 (19.24%) of MDI prescriptions. Carbon emissions from ICS MDIs and triple therapy MDIs made up only a small proportion of emissions at 9.7 tonnes (3.30%) and 7.6 tonnes (2.58%), respectively. MDI prescriptions generated nearly 99% of total carbon emissions, with the highest-emission devices producing over 60 times more CO^2^-equivalent emissions than the lowest-emission alternatives.

**Table 2:** Device-Specific Carbon Emissions Profile.

| Medication Class | Device Type | Device Examples | Carbon Emissions per Device (grams CO<sub>2</sub>e) | Total Prescriptions | % of Total Prescriptions | Total Emissions (tonnes) | % of Total Emissions |
| --- | --- | --- | --- | --- | --- | --- | --- |
| SABA | MDI | Ventolin, Airomir, | 28,700 | 7,534 | 44.91% | 190.3 | 64.22% |
| SABA | DPI | Terbutaline Turbuhaler | 790 | 76 | 0.45% | 0.06 | 0.02% |
| SAMA | MDI | Atrovent | 14,590 | 1,235 | 7.36% | 18.0 | 6.08% |
| LABA | DPI | Serevent Diskus | 790 | 26 | 0.16% | 0.02 | 0.01% |
| LAMA | DPI | Spiriva HandiHaler | 790 | 717 | 4.28% | 0.57 | 0.19% |
| LAMA | SMI | Spiriva Respimat | 775 | 774 | 4.62% | 0.60 | 0.20% |
| ICS | MDI | Flixotide, Alvesco | 20,000 | 484 | 2.89% | 9.7 | 3.27% |
| ICS | DPI | Pulmicort Turbuhaler | 790 | 272 | 1.62% | 0.21 | 0.07% |
| LABA/LAMA | DPI | Anoro Ellipta | 790 | 179 | 1.07% | 0.14 | 0.05% |
| LABA/LAMA | SMI | Stiolto Respimat | 775 | 235 | 1.40% | 0.18 | 0.06% |
| ICS/LABA | MDI | Symbicort Rapihaler, Flutiform | 36,500 | 2,319 | 13.83% | 67.2 | 22.67% |
| ICS/LABA | DPI | Symbicort Turbuhaler, Seretide Accuhaler | 790 | 1,464 | 8.73% | 1.16 | 0.39% |
| ICS/LABA/LAMA | MDI | Trimbow | 15,000 | 480 | 2.86% | 7.6 | 2.56% |
| ICS/LABA/LAMA | DPI | Trelegy Ellipta | 790 | 974 | 5.81% | 0.77 | 0.26% |
| **TOTAL** | **All Devices** | — | — | **16,769** | **100.00%** | **296.35** | **100.00%** |

**Legend:** SABA = Short-Acting Beta-Agonist; SAMA = Short-Acting Muscarinic Antagonist; LABA = Long-Acting Beta-Agonist; LAMA = Long-Acting Muscarinic Antagonist; ICS = Inhaled Corticosteroid; MDI = Metered Dose Inhaler; DPI = Dry Powder Inhaler; SMI = Soft Mist Inhaler; CO<sub>2</sub>e = Carbon Dioxide Equivalent. Carbon emissions data based on lifecycle assessment including manufacturing, use, and disposal phases. Device examples represent commonly prescribed brands available on the Australian Pharmaceutical Benefits Scheme.

**Table 3:** Current Carbon Emissions by Device Type.

| Device Type | Current Emissions (tonnes) | Percentage of Total |
| --- | --- | --- |
| Metered Dose Inhalers (MDI) | 292.77 | 98.79% |
| Dry Powder Inhalers (DPI) | 2.79 | 0.94% |
| Soft Mist Inhalers (SMI) | 0.78 | 0.26% |
| **TOTAL** | **296.35** | **100.00%** |

**Legend:** MDI = Metered Dose Inhaler; DPI = Dry Powder Inhaler; SMI = Soft Mist Inhaler.

To assess the potential for carbon reduction, we evaluated the environmental impact of switching MDIs to DPI alternatives where clinically appropriate as outlines in Table 4. A suitable DPI alternative was identified for each MDI device, and the carbon emission savings were calculated. Each DPI device has been previously calculated to have carbon emissions of less than 900g of carbon, compared to 14,279 – 36,500g for the MDI devices prescribed. No SAMA agent is available in DPI or SMI form, so no reduction in carbon emissions could be calculated for this class. For each medication class where either an MDI or DPI could be prescribed, the MDI’s carbon footprint, on average, was 34.75 times greater per device than that of the DPI.

**Table 4:**
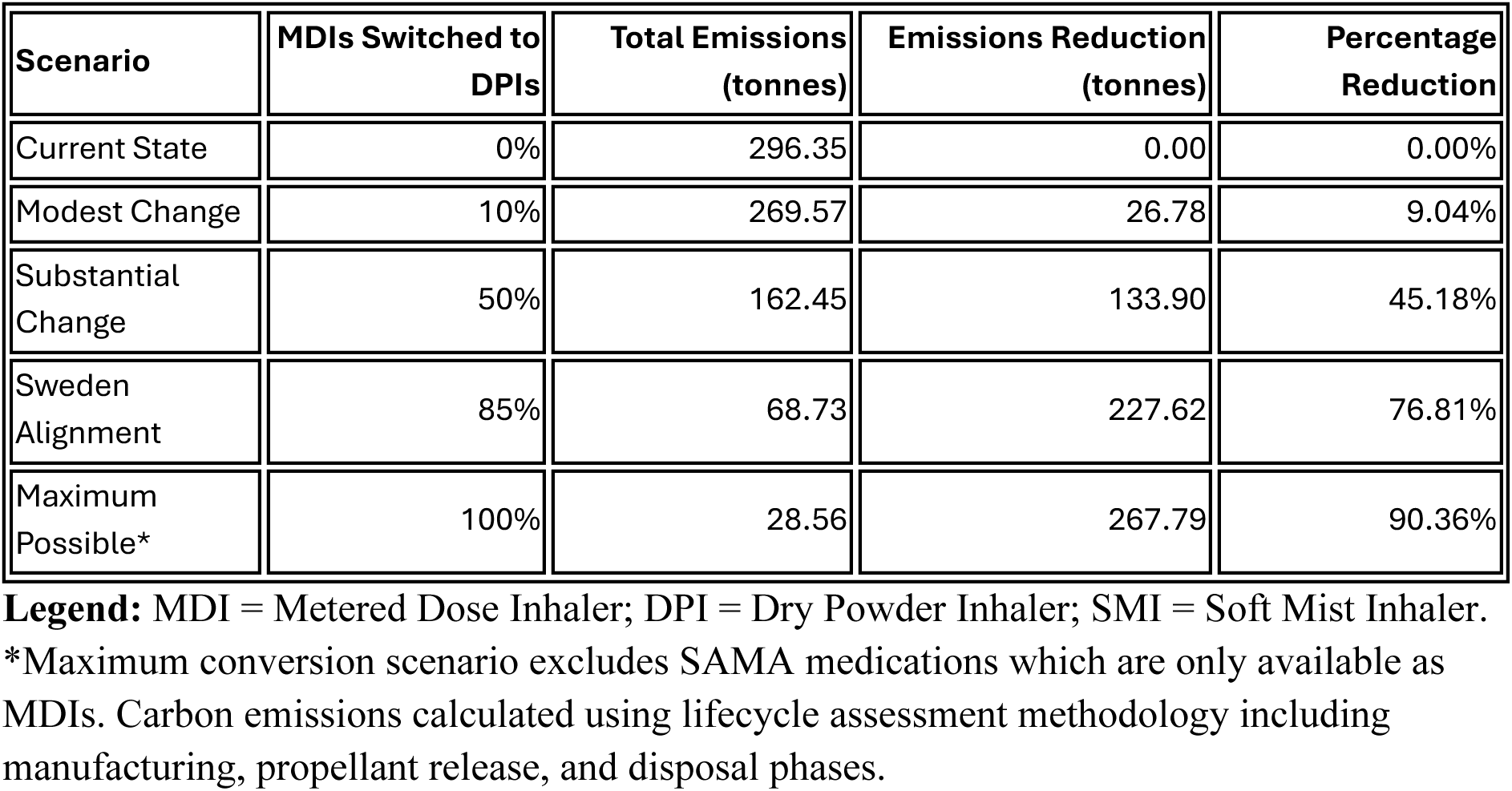
Carbon Reduction Potential by Switching MDIs to DPIs.

The potential carbon savings from device switching were substantial as demonstrated in Table 4. If 100% of MDIs were replaced with DPIs where possible, 267.79 tonnes of carbon would be reduced, with a 90.36% reduction in total carbon emissions. This is the equivalent of 1,479,498 km of automobile driving or driving 3.5 times around the Earth. Of the remaining 28.35 tonnes of carbon emissions, most would be from SAMA prescriptions (18.0 tonnes, 63.09%) and SABAs (4.57 tonnes, 15.99%).] More modest changes would still yield significant benefits according to our modelling scenarios (Table 4): an 85% change to DPIs would result in a 227.62 tonnes reduction (76.81% reduction), a 50% change would result in a 133.89-tonne reduction (45.18%), and even a 10% change would result in a 26.78-tonne reduction (9.05%).

## Discussion

This study demonstrates that the inhaler discharge prescriptions at Western Health comprise largely metered-dose inhaler (MDI) prescriptions, which make up the vast majority of the carbon emissions (71.87% of prescriptions generating 98.79% of emissions as shown in Tables 1 and 3). Although there is minimal published data on hospital discharge prescriptions in Australia, this data is consistent with prior Australian pharmacy sales data and with international prescriptions (4), and with recent local evidence from the nearby Royal Melbourne Hospital, which demonstrated similarly high MDI prescribing rates (79% of 9,246 inhalers) (8). The potential for substantial change is demonstrated by Sweden’s healthcare system, which prescribes only 13% of inhalers as MDIs (6), compared to Australia’s and the United Kingdom’s approximate 70%(2), achieving this without compromising asthma or COPD outcomes. In the UK, the National Health Service (NHS) has committed to net-zero carbon emissions by 2040, with 3% of the NHS’s current carbon emissions coming from inhalers (11), while in Australia, the National Health and Climate Strategy identifies inhaler emission reduction as an improvement area (7).

While our findings demonstrate substantial potential for carbon reduction through device switching, clinical appropriateness must remain the primary consideration. MDIs with spacer devices may be preferred in certain clinical scenarios, including young children under 6 years of age who may not generate sufficient inspiratory flow for DPIs, patients with severe COPD exacerbations with compromised inspiratory flow rates, and emergency situations where MDIs provide more reliable drug delivery regardless of patient effort (12). However, our finding that triple therapy inhalers are disproportionately prescribed as DPIs (67% vs 33% MDIs) in patients with severe lung disease suggests that DPIs are clinically appropriate even for severely unwell cohorts. The variability in device prescriptions between medication classes suggests that prescriber familiarity and institutional habits significantly impact device choice beyond clinical factors alone. Given that many Scandinavian countries and Japan prescribe a large proportion of inhalers as DPIs without compromising safety (4), a higher proportion of inhalers could likely be safely prescribed in DPI form.

Beyond device selection, the types of medications prescribed also present opportunities for both clinical and environmental improvement. The predominance of SABA prescriptions (45.38% of all inhalers) represents both an environmental and clinical concern. Recent Australian and international asthma guidelines have recommended first-line therapy of combined preventer and reliever inhalers instead of short-acting beta-agonists. Both the Asthma Handbook Australia and the Global Initiative for Asthma guidelines recommend ICS/ LABA anti-inflammatory reliever (AIR) therapy as the first-line treatment option for most adults with asthma (13)(14). This guideline shift is supported by evidence that short-acting inhaler use is associated with higher mortality and hospitalisation compared to longer-acting agents due to tachyphylaxis and inability to treat underlying inflammation 13/7/2025 2:26:00 pm. A shift from short-acting agents to preventers would therefore benefit both patient care and the environment.

This study demonstrates that strategic changes in prescription habits could substantially reduce healthcare carbon emissions. Almost the entire carbon footprint from inhalers prescribed at Western Health comes from MDIs (98.79%), which would be almost entirely eliminated if a substantial portion were changed to DPIs. A change to 85% DPI prescriptions in line with Swedish prescriptions would result in a 76.81% reduction in carbon emissions. These findings align with the TSANZ position statement’s emphasis that “the carbon footprint of DPI and SMI devices can be 100-200-fold lower than those of pMDIs”, providing local Australian hospital data to support this international evidence (2). Hospital discharge prescribing represents a particularly important intervention point as discharge prescriptions frequently become templates for ongoing community prescribing, creating multiplier effects for environmental benefits.

However, implementing such changes requires careful consideration of multiple factors. Caution is required with changes to inhaler prescriptions. Prior schemes that have changed patients to dry powder inhalers without substantial patient consultation have met with significant backlash and have resulted in worsened disease control (15). Furthermore, patients may have strong device preferences, and DPIs are typically more expensive than MDIs, creating both clinical and economic considerations. There has also been feedback from patient advocacy groups discussing the environmental impact of inhalers, which can be perceived as shifting responsibility away from international pharmaceutical corporations and the government and inappropriately transferring responsibility to patients (16). Individual changes should involve close patient consultation, with systemic changes requiring stakeholder involvement, including patient advocacy groups, healthcare providers, and policymakers.

Several methodological limitations of this study should also be acknowledged. Our analysis examined prescriptions rather than actual dispensing data, assuming each prescription was filled once without accounting for partial filling or non-adherence. However, the environmental impact of propellants occurs regardless of medication usage, as hydrofluorocarbons eventually escape into the atmosphere when devices deteriorate. The carbon footprint calculations rely on estimates from Tirumalasetty et al., which carry uncertainty due to reliance on material safety data sheets rather than direct measurement (9). A significant limitation is the absence of clinical data on patient characteristics, disease severity, and appropriateness of device selection. Without access to patient records, we could not assess whether MDI prescriptions were clinically indicated (e.g., for patients with poor inspiratory capacity, young children, or acute exacerbations) or represented missed opportunities for environmentally preferable alternatives. Additionally, certain med,ications such as ipratropium (SAMA), are only available as MDIs, limiting the potential for complete elimination of high-carbon devices. While DPIs may be more expensive than MDIs for the healthcare system, a comprehensive cost-benefit analysis was beyond the scope of this study but would be valuable for future policy implementation.

Looking forward, several research priorities emerge from these findings. Future research should investigate patient and clinician attitudes toward device switching, conduct cost-effectiveness analyses that incorporate environmental and economic factors, and monitor long-term clinical outcomes and adherence patterns following device switches. As Australia develops its National Health and Climate Strategy, this study provides evidence that strategic changes in hospital inhaler prescribing could substantially reduce healthcare greenhouse gas emissions while maintaining quality patient care

## Data Availability

All data produced in the present study are available upon reasonable request to the authors.

## List of abbreviations

CO₂: Carbon dioxide
CO₂e: Carbon dioxide equivalent
COPD: Chronic obstructive pulmonary disease
DPI: Dry powder inhaler
GWP: Global warming potential
HFC: Hydrofluorocarbon
ICS: Inhaled corticosteroid
ICS/LABA: Inhaled corticosteroid/Long-acting beta-agonist combination
ICS/LABA/LAMA: Inhaled corticosteroid/Long-acting beta-agonist/Long-acting muscarinic antagonist triple therapy
LABA: Long-acting beta-agonist
LAMA: Long-acting muscarinic antagonist
LABA/LAMA: Long-acting beta-agonist/Long-acting muscarinic antagonist combination
MDI: Metered dose inhaler
NHS: National Health Service
SABA: Short-acting beta-agonist
SAMA: Short-acting muscarinic antagonist
SMI: Soft mist inhaler
TSANZ: Thoracic Society of Australia and New Zealand
UK: United Kingdom

## Declarations

### Ethics approval

This study was approved by the Western Health Research Office as a Minimal Risk Research project (QA/114629/WH-2025-485634(v3); Local Reference Number: QA2024.105) on 3 June 2025. The project met the requirements of the National Statement on Ethical Conduct in Human Research (NHMRC, 2023), the Australian Code for the Responsible Conduct of Research (NHMRC, 2018), and was consistent with Ethical Considerations in Quality Assurance and Evaluation Activities (NHMRC, 2014). As this was a retrospective study using de-identified prescription data extracted from electronic medical records, individual patient consent was not required.

### Consent for publication

Not applicable

### Availability of data and materials

The datasets generated and analysed during this study are not publicly available due to patient confidentiality restrictions and the terms of ethics approval but aggregate results are presented in full within this article.

### Competing interests

J.T. has no competing interests to declare

L.J. has no competing interests to declare

### Funding

None received

### Authors’ contributions

J.T. conceived and designed the study, obtained ethics approval, performed data extraction and analysis, conducted all carbon emission calculations, and drafted the manuscript.

L.J. contributed to study design, assisted with manuscript writing, and provided critical revision of the manuscript for important intellectual content. Both authors read and approved the final manuscript.

## Acknowledgements

Not applicable

## Notes

### Competing Interest Statement

The authors have declared no competing interest.

### Funding Statement

This study did not receive any funding

### Author Declarations

Western Health Ethics Committee of Western Health gave ethical approval for this work (Ethics number 7326409). Per the Western Health Ethics Committee "The proposal meets the requirements of the National Statement on Ethical Conduct in Human Research (NHMRC, 2023) and the Australian Code for the Responsible Conduct of Research (NHMRC, 2018); it is consistent with Ethical Considerations in Quality Assurance and Evaluation Activities (NHMRC, 2014). The project is authorised to commence at Western Health."

